# International Classification of Diseases (ICD) Codes for Congenital Heart Defects (CHD) Have Variable and Limited Accuracy for Detecting CHD Cases

**DOI:** 10.1101/2023.04.20.23288898

**Authors:** Lindsey C. Ivey, Fred H. Rodriguez, Haoming Shi, Cohen Chong, Joy Chen, Cheryl Raskind-Hood, Karrie F. Downing, Sherry L. Farr, Wendy M. Book

**Affiliations:** Division of Cardiology, Emory University School of Medicine, Division of Cardiology, Atlanta, GA; Children’s Healthcare of Atlanta Cardiology, Atlanta, GA; Department of Biomedical Engineering, Georgia Institute of Technology and Emory University, Atlanta, GA; Emory University Rollins School of Public Health, Atlanta, GA; PicnicHealth, San Francisco, CA; National Center on Birth Defects and Developmental Disabilities, Centers for Disease Control and Prevention, Atlanta, GA

**Keywords:** Congenital heart defects, outcomes research, epidemiology

## Abstract

**Background:** Administrative data permits analysis of large cohorts but relies on International Classification of Diseases, Ninth and Tenth Revision, Clinical Modification (ICD) codes that may not reflect true congenital heart defects (CHD).

**Methods:** 1497 cases with at least one encounter between 1/1/2010 – 12/31/2019 in two healthcare systems (one adult, one pediatric) identified by at least one of 87 ICD CHD codes were validated through chart review for the presence of CHD and CHD anatomic group.

**Results:** Inter- and intra-observer reliability averaged > 95%. Positive predictive value (PPV) of ICD codes for CHD was 68.1% (1020/1497) overall, 94.6% (123/130) for cases identified in both healthcare systems, 95.8% (249/260) for severe codes, 52.6% (370/703) for shunt codes, 75.9% (243/320) for valve codes, 73.5% (119/162) for shunt and valve codes, and 75.0% (39/52) for “Other CHD” (7 ICD codes). PPV for cases with >1 unique CHD code was 85.4% (503/589) vs. 56.3% (498/884) for one CHD code. Of cases with secundum atrial septal defect ICD codes 745.5/Q21.1 in isolation, 30.9% (123/398) had a confirmed CHD. Patent foramen ovale was present in 66.2% (316/477) of false positives (FP). The median number of unique CHD-coded encounters was higher for true positives (TP) than FP (2.0; interquartile range [IQR]: 1.0-3.0 vs 1.0; IQR:1.0-1.0, respectively, p<0.0001). TP had younger mean age at first encounter with a CHD code than FP (22.4 years vs 26.3 years, p=0.0017).

**Conclusion:** PPV of CHD ICD codes varies by characteristics for detection of CHD by ICD code and anatomic grouping. While an ICD code for severe CHD and/or the presence of a case in more than one data source, regardless of anatomic group, is associated with higher PPV for CHD, most TP cases did not have these characteristics. The development of algorithms to improve accuracy may improve administrative data for CHD surveillance.

## INTRODUCTION

The prevalence of congenital heart defects (CHD) beyond the first year of life varies across the United States depending on the data source, case definition, and methodology used by investigators^1-3^. Public health surveillance to determine disease prevalence and long-term outcomes for this population often relies on large administrative and clinical datasets from various sources including electronic medical records (eMR) and billing data from hospital systems and government agencies^4-6^. The reliability of CHD case detection in eMR and administrative data depends on the accuracy of International Classification of Diseases, Ninth and Tenth Revision, Clinical Modification (ICD-9-CM & ICD-10-CM) diagnosis code documentation in patient records^7-9^. Identifying CHD cases by ICD codes may be prone to errors from misspecification, upcoding, rule-out codes, and other issues that contribute to false positive (FP) and false negative classifications^7^. CHD studies often use ICD-9-CM and ICD-10-CM code groups, 745.xx – 747.xx (63 codes) and Q20.x – Q26.x (73 codes) respectively, to define a CHD cohort^2, 10^. Some of the codes in this group, however, represent gastrointestinal, renal, spinal, extremity, umbilical artery, and other non-cardiac vascular anomalies. ICD-10-CM relocated many non-cardiac vascular anomalies into groups Q27.x and Q28.x, but non-specific CHD codes remain within Q20.x – Q26.x. Additionally, some codes intended to represent CHD lack diagnostic specificity and have resulted in poor positive predictive values (PPV) for true CHD identification^2, 9, 11-15^. For example, ICD-9-CM code 745.5, which represents both secundum atrial septal defect (ASD) and patent foramen ovale (PFO) had a PPV of only 23.7% in an adolescent and adult population^2, 11^. Exclusion of code 745.5 and CHD codes categorized as “other” increased PPV of CHD codes over 10 percentage points to 86.5%,^16^ but at the expense of losing an estimated 24% of true CHD cases with code 745.5 in an administrative data set.

Differences in coding practices between administrative and clinical databases can lead to differences in case ascertainment, and subsequently, prevalence and outcome estimations^17, 18^. A hierarchal algorithm of 17 CHD subgroups found that the c-statistic for the code hierarchy alone was 0.79 (0.77–0.80) and improved to 0.89 (0.88–0.90) after age, encounter type, and provider type were included in the algorithm^9, 19^. Therefore, algorithms and hierarchies^2, 19-21^ accounting for limitations of administrative coding are often applied to restrict a cohort to those individuals most likely to have the condition of interest, at the expense of unintentional exclusion of true cases from the dataset.

Due to limitations in administrative data, restrictive case definitions may underestimate patients with CHD; however, unrestrictive case definitions that include the broader code groups 745.xx – 747.xx and Q20.x – Q26.x may overestimate patients with CHD. To understand limitations of and improve the quality of administrative data for CHD surveillance, we evaluated the PPV of select ICD-9-CM and ICD-10-CM CHD codes in two data sources and compared features associated with both true positive (TP) and FP cases.

## METHODS

This study utilized data from two Georgia-based statewide multi-hospital and outpatient tertiary healthcare systems — a pediatric healthcare system (PHS), and an adult healthcare system (AHS). Data were collected under a Cooperative Agreement between Emory University and Centers for Disease Control and Prevention (CDC-RFA-DD19-1202B). Study approval by the Emory University Institutional Review Board (IRB) was granted on August 26, 2020 (IRB# STUDY00001030) and included a complete waiver of HIPAA authorization as well as a waiver of informed consent.

### Case Selection for Chart Abstraction

The cohort identified for case validation consisted of individuals born between 1/1/1955 and 12/31/2019, who were identified in at least one of the two clinical data sources with a healthcare encounter between 1/1/2010–12/31/2019 and with at least one of 87 selected CHD ICD-9-CM and ICD-10-CM codes noted in Table 1. Forty-nine ICD codes in the CHD code groups 745.xx – 747.xx and Q20.x – Q26.x (25 ICD-9-CM and 24 ICD-10-CM) were excluded due to lack of specificity for CHD. We hereafter referred to those meeting the case definition as “cases” even if determined not to have a CHD after validation. Because patent ductus arteriosus (PDA) can naturally resolve in infancy, cases with a PDA code associated only with encounters in the first three months of life were excluded from selection, unless they had other included CHD codes. From a cohort of 15,504 individuals identified in the AHS meeting inclusion criteria, 750 cases at least 18 years of age as of 1/1/2010 were randomly selected. From a cohort of 36,399 in the PHS who met inclusion criteria, another 750 cases were randomly selected for chart abstraction. Encounter level CHD codes were grouped based on native anatomy into severe, shunt and valve, only shunt, only valve, and other CHD^2^ (Table 1).

**Table 1:**
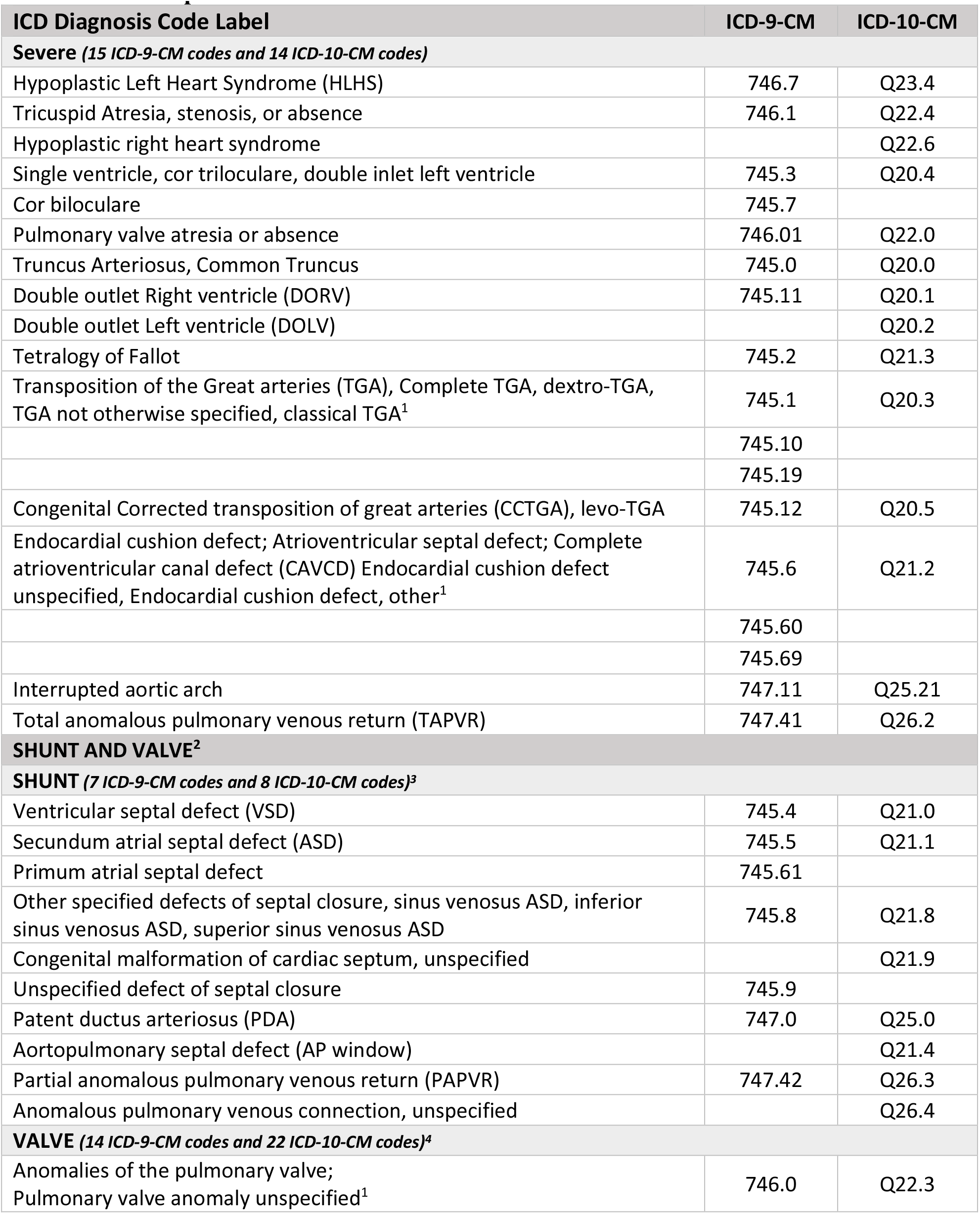
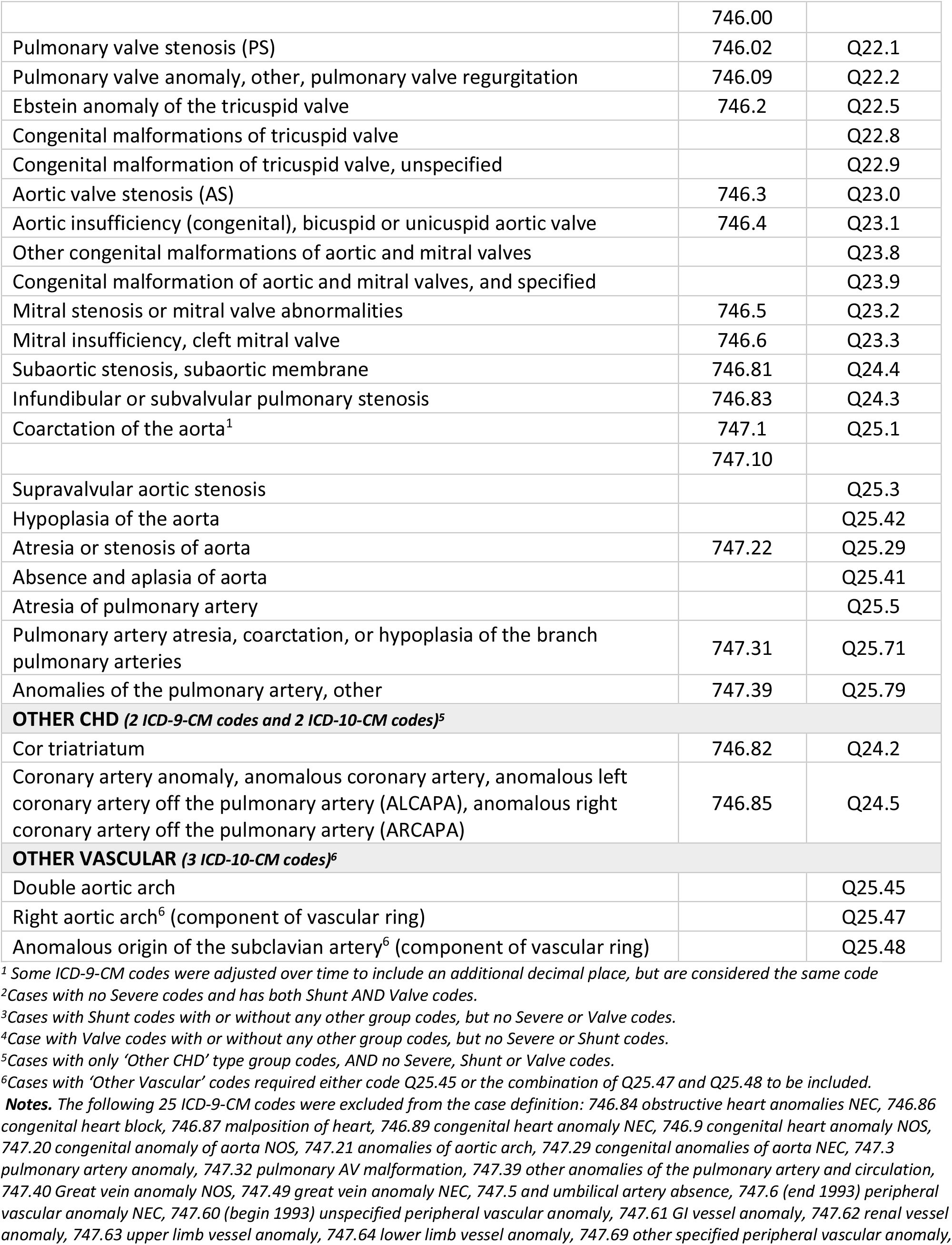
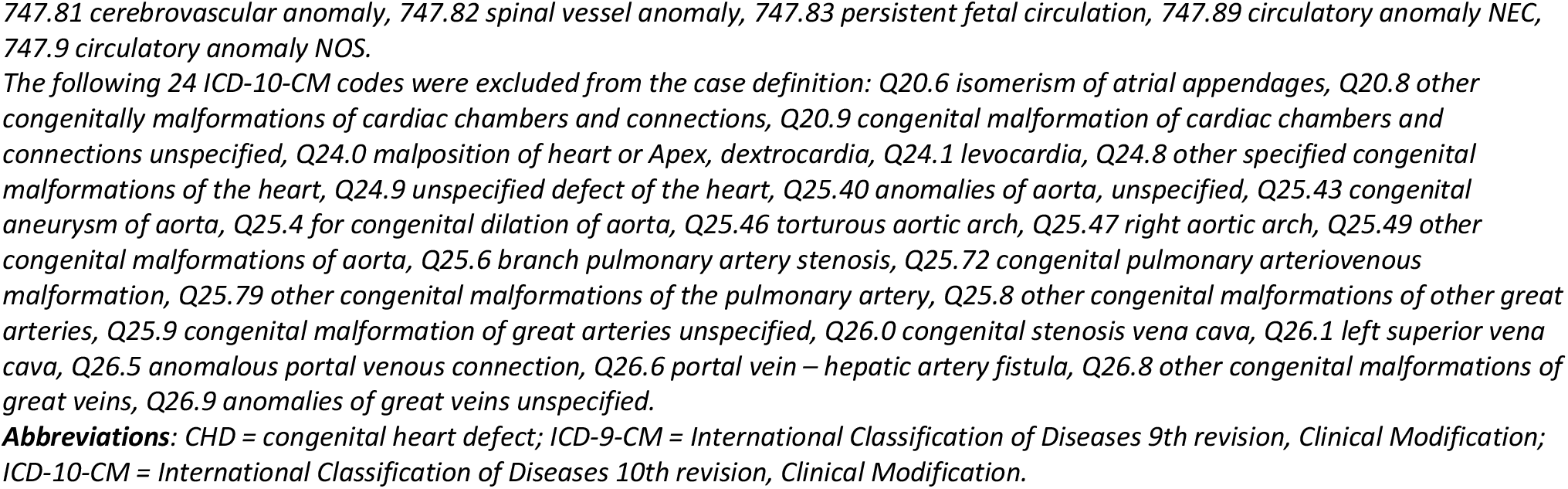
ICD-9-CM and ICD-10-CM CHD Code Inclusion Criteria for the Cohort from which the Random Sample was Selected.

### Research Electronic Data Capture (REDCap) Database Development

Study data were collected and managed using Research Electronic Data Capture (REDCap), a secure, web-based application designed to support data capture for research studies.^22, 23^ Only demographics were entered into the REDCap database to enable abstractors to locate the eMRs for each case, with abstractors blinded to ICD codes. The REDCap database was developed, tested, and revised using an existing dataset from the same facilities, with clinician input.

### Abstractor Training, Process, and Quality Review

Two board certified CHD clinicians (WMB and FHR) trained the abstractors through an iterative process to identify the correct CHD diagnosis for a case. The clinicians reviewed cases weekly for accuracy. Inter- and intra-observer reliability between chart abstractors and CHD clinicians were measured in three formal training rounds for 102 test cases, in an iterative process using the 2010–2019 dataset. The same cases were randomly assigned to each abstractor and clinician. Clinician determination based on chart abstraction was considered the gold standard. For the training cases, inter-observer and intra-observer reliability averaged 95%. For each case, abstractors determined ***CHD Yes/No/Unknown*** and the anatomic group based on review of text clinical documents of any available records, including healthcare system-associated health network records and scanned outside records. ***Unknown CHD*** was defined as the absence of clinical records sufficient to determine if a case has a CHD, such as no encounter notes in the eMR. ***Not CHD*** was defined in the medical record documentation as the absence of heart defects recorded in imaging or provider notes. PFO, and other non-CHD cardiac diagnoses were noted for all ***Not CHD*** and reviewed by clinicians. During the validation process, a PDA in isolation in an infant less than 3 months of age was not considered a CHD unless the PDA persisted after this age. Similarly, peripheral branch pulmonary stenosis was noted in pediatric cases, but not considered a CHD in infancy in the absence of other associated heart defects. All ***Not CHD*** and ***Unknown CHD*** cases were reviewed by at least three abstractors and two board certified CHD clinicians, to make a final determination. For cases with ***Unknown CHD*** status that remained unknown after review, the cases were replaced with other randomly selected cases.

### Chart Abstraction and Case Validation

Abstractors and clinicians reviewed 1500 cases meeting criteria for case selection, not including the training cases and 19 ***Unknown CHD*** cases that were replaced (three cases had to be excluded due to being pulled using a code that should have been excluded in isolation, ICD-9-CM 747.3 and ICD-10-CM Q25.6), creating a final data set of 1497 cases. We categorized cases into six anatomic groups and developed rules to best determine when multiple conflicting groups were possible. The CHD diagnosis was based on review of healthcare clinical notes, imaging notes, operative reports, and scanned clinical notes from outside each institution. Billing ICD codes that are sometimes found in the eMR were not used to confirm a diagnosis. As cases could potentially have records in both PHS and AHS, chart abstractors reviewed medical records from both healthcare systems during the case validation process. Cases flagged to review with the clinicians included all cases with atrioventricular canal defects (partial, complete, or transitional), transposition of the great arteries (TGA) (either dextro-TGA or levo-TGA), cases classified as shunt + valve, ***Unknown CHD, Not CHD***, or those with conflicting diagnoses. Initially, clinicians reviewed all ***Not CHD, Yes PFO*** cases. Once consistency and accuracy were assured, the clinicians no longer reviewed cases clearly understood to be PFOs by the abstractor. ***Unknown CHD*** cases with no medical information were linked back to the data source to verify the presence of a CHD ICD code in the medical record as a quality check. Forty percent of cases (n=593) underwent adjudication by two clinicians.

Additionally, female cases between 11–50 years-old at any time in the surveillance window were checked for the presence of a pregnancy in the eMR and recorded in REDCap due to the possibility that a CHD code associated with a pregnant women’s chart might reflect CHD in the fetus.

### Intra- and Inter-observer Reliability

To monitor intra-observer reliability, 5% of each abstractor’s cases were randomly duplicated and reviewed. To monitor inter-observer reliability throughout the study, 10% of cases were randomly duplicated and assigned to the clinicians, regardless of initial abstractor assignment. Intra-observer reliability ranged from 95–100% and inter-observer reliability ranged from 88–100% (Table 2).

**Table 2.** Intra-observer and Inter-observer Reliability Results for Randomly Selected Cases from the 1500 Validated Dataset.

| Observer | CHD<br>Yes or No<br>Agreement | Percent<br>Agreement |
| --- | --- | --- |
| **Intra-Observer Reliability<sup>1</sup>** |  |  |
| Abstractor1 | 25/25 | 100% |
| Abstractor2 | 20/21 | 95% |
| Abstractor3 | 21/22 | 95% |
| Abstractor4<sup>2</sup> | 7/7 | 100% |
| **Inter-Observer Reliability<sup>3</sup>** |  |  |
| Clinician1<sup>4</sup> vs Abstractor1 | 23/25 | 92% |
| Clinician1 vs Abstractor2 | 27/27 | 100% |
| Clinician1 vs Abstractor3 | 21/24 | 88% |
| Clinician2<sup>5</sup> vs Abstractor1 | 25/27 | 93% |
| Clinician2 vs Abstractor2 | 20/21 | 95% |
| Clinician2 vs Abstractor3 | 15/15 | 100% |
| Clinician2 vs Abstractor4 | 12/12 | 100% |

<sup>1</sup>5% of each abstractor's cases were randomly duplicated and validated again by the same abstractor to assess intra-observer reliability.
<sup>2</sup>Abstractor 4 only abstracted cases from PHS
<sup>3</sup>10% of validated cases were randomly duplicated and validated by two different clinicians to assess inter-observer reliability.
<sup>4</sup>Clinician 1 abstracted cases from AHS
<sup>5</sup>Clinician 2 abstracted cases from PHS
AHS= Adult Healthcare System, PHS=Pediatric Healthcare System

### Statistical Analysis

In addition to data source, sex, and anatomic severity groups, we assessed encounter characteristics including the number of unique CHD ICD codes documented per case, number of unique encounters with a CHD ICD code documented per case, number of cardiac encounters per case, number of imaging encounters apart from a cardiac encounter per case, and number of non-cardiac encounters per case. Cardiac encounters were defined as either cardiac encounter type, an encounter with a cardiologist provider type, or an encounter with a National Provider Identifier (NPI) number with a cardiac specialty. Imaging encounters were defined as Current Procedural Terminology (CPT) codes for cardiac imaging (echocardiograms, cardiac CT scans, and cardiac MRIs; eAppendix A) and did not include those that were also a cardiac encounter. Non-cardiac encounters were those that did not fall within the cardiac or imaging encounter definition. Additionally, age in years at first qualifying encounter (FQE, defined as the first encounter with a CHD code in the surveillance period) was collected for each case.

Percentages by data source, sex, anatomic severity groups, and median encounter characteristics were compared for patients with true CHD and patients without CHD (FP, i.e., cases determined not to have a CHD in chart abstraction) as well as for FP with PFO and without PFO using Pearson’s chi-square test of independence for the categorical characteristics, and the non-parametric two-sample Kolmogorov-Smirnov test for encounter characteristics (chosen due to heavily skewed distributions). In addition, group differences in median age in years at the FQE were analyzed via t-test. PPV was calculated as percent of TP: (TP) / [TP + FP]. Statistical significance was defined as p-value < 0.05.

## RESULTS

Of the 1497 included validated cases with at least one of the 87 codes in ICD-9-CM code group 745.xx – 747.xx and ICD-10-CM code group Q20.x – Q26.x (Table 1), 68.1% (n=1020) were TP and 31.9% (n=477) were FP. Of the 1020 TP, 24.4% (n=249) had severe CHD, 11.7% (n=119) had shunt and valve CHD, 36.3% (n=370) had shunt CHD, 23.8% (n=243) had valve CHD, and 3.8% (n=39) had “other CHD” or “other vascular” CHD. Among FP, 66.2% (316/477) were confirmed to have a PFO. Of FP confirmed to have neither a CHD nor a PFO, 42.2% (68/161) had other cardiac diagnoses, consisting of heritable and metabolic cardiomyopathies (n= 20), coronary artery disease (n=14), non-congenital valvular heart disease (n=13), arrythmias (n=11), heritable aortopathies (n=5), pericardial disease (n=2), dextrocardia (n=1) and other cardiac diagnoses (n=2).

In Table 3, PPV of CHD ICD-9-CM and ICD-10-CM codes was 64.8% (481/742) in AHS data, 66.6% (416/625) in PHS data, and 94.6% (123/130) for those individuals who had clinical notes in both the AHS and PHS data. Males were as likely to be TP as females (p=0.2525).

**Table 3.**
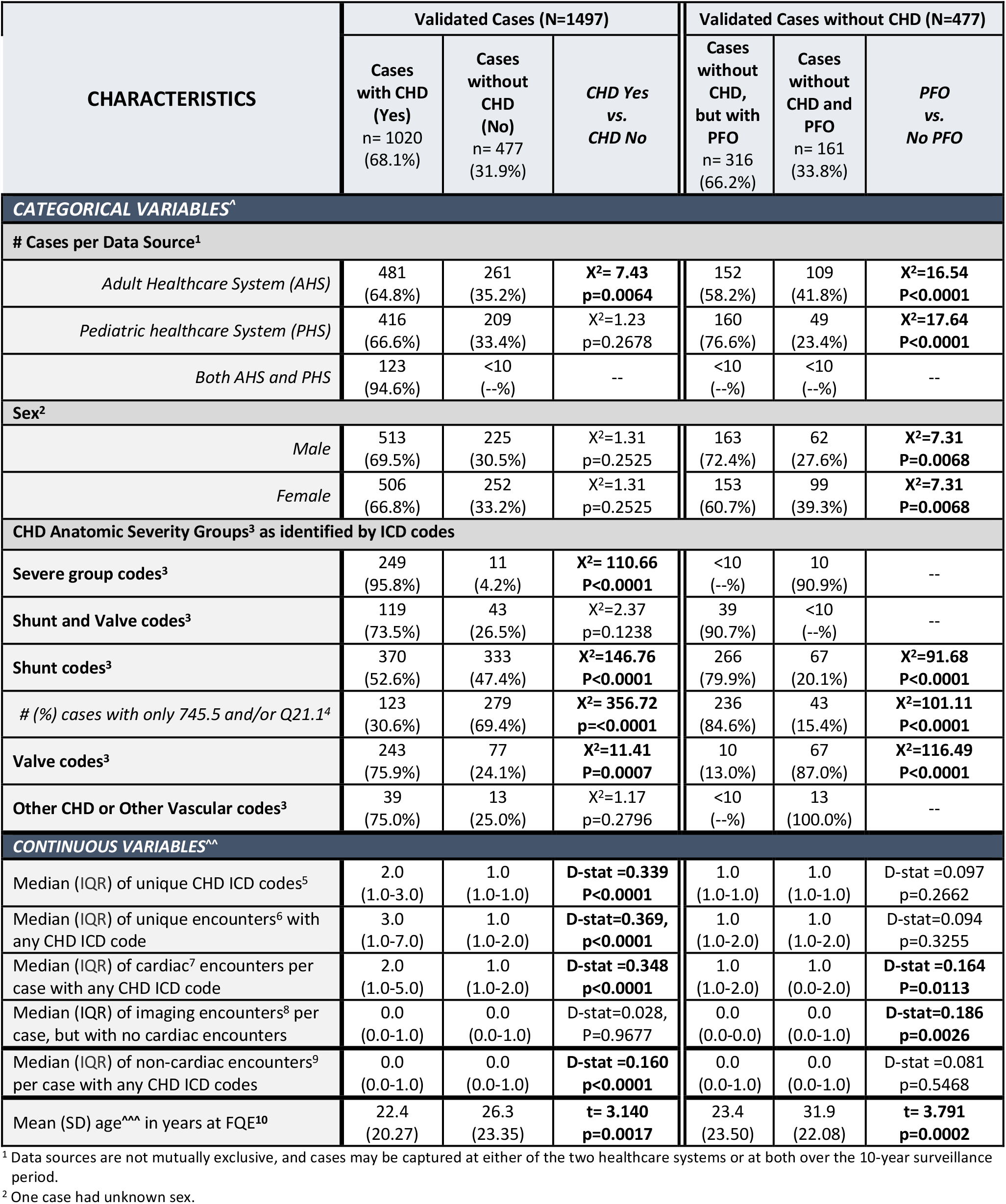
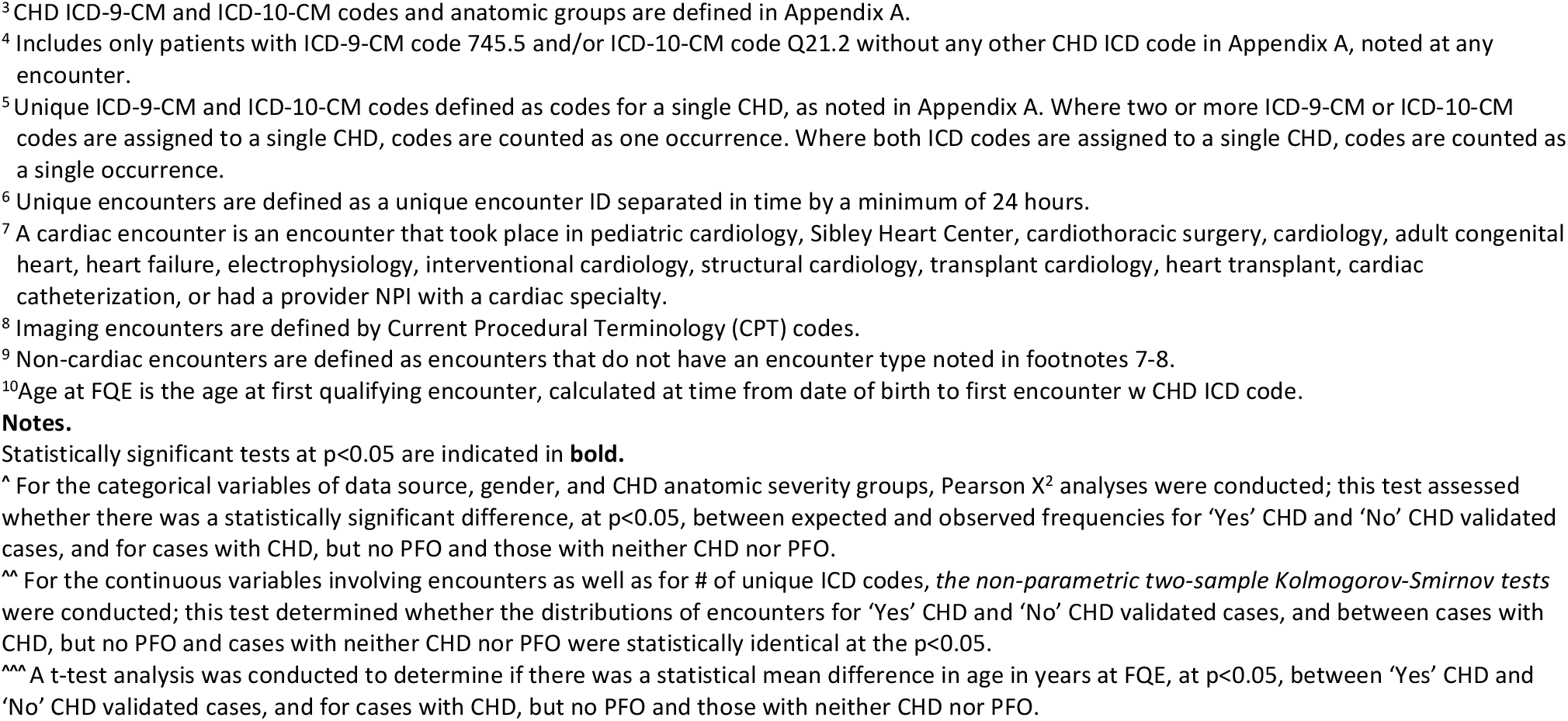
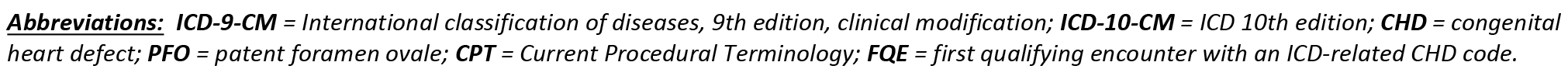
Characteristics of 1497 Validated Cases with at least one CHD ICD-9-CM and/or ICD-10-CM code(s)^1^ and with at least one encounter between 1/1/2010 -12/31/2019 in an Adult Healthcare System and a Pediatric Healthcare System.

Overall, the PPV for CHD among cases was 95.8% for severe codes (249/260), 73.5% (119/162) for shunt and valve codes, 52.6% (370/703) for shunt codes, 75.9% (243/320) for valve codes, and 75.0% (39/52) for “Other CHD” or “Other vascular” codes (Table 3). Just over a quarter of the cohort, 26.9% (402/1497), had only 745.5 and/or Q21.1 (secundum ASD) shunt codes without other CHD-related codes. Of these, 30.6% (123/402) were TP and 69.4% (279/402) were FP. Overall, TP with only secundum ASD codes represented 12.1% (123/1020) of all TP.

The distribution of the number of unique CHD codes was significantly different for TP than FP (Median: 2.0; interquartile range [IQR]: 1.0–3.0 vs. Median: 1.0; IQR:1.0–1.0, p<0.0001) (Table 3). The distribution of unique encounters with CHD codes was also significantly higher (p<0.0001) for TP (3.0; IQR: 1.0–7.0) than FP (1.0; IQR:1.0–2.0). There was a significant difference between the distribution of cardiac encounters for TP (2.0; IQR:1.0–5.0) and FP (1.0; IQR: 1.0–2.0), p<0.0001. TP cases had a younger mean age at FQE than FP (22.4 years vs 26.3 years, p=0.0017).

Among FP, males were more likely to have a PFO than females (72.4% vs 60.7%, p=0.0068). FP cases with codes in the shunt group, and specifically isolated 745.5 and/or Q21.1 codes, were more likely to have a PFO, whereas FP with codes in the valve group were less likely to have a PFO (all p<0.0001) (Table 3). The median number of unique CHD ICD codes was similar between FP cases with PFO and FP cases without PFO (both medians=1.0, p=0.2662). FP cases with PFO did not have any differences in the distributions of unique encounters or non-cardiac encounters. The mean age at FQE of FP cases with a PFO was lower than for FP cases without a PFO (23.4 years vs 31.9 years, p<0.0001).

Table 4 shows PPV of the CHD-related ICD codes for individuals identified in AHS only (n=742), PHS only (n=602), in both AHS and PHS (n=129), and among all cases with CHD coded encounters in the surveillance period (i.e., AHS and/or PHS combined; n=1473). The presence of more than one unique CHD ICD code resulted in higher PPV compared to those with only one unique CHD code (85.4% vs. 56.3%, respectively; p<0.0001). When assessing PPV stratified by date range, overall PPV differed slightly between 1/1/2010 to 9/30/2015 (ICD-9-CM coding era) (75.3%; 579/769) and 10/1/2015 to 12/31/2019 (ICD-10-CM coding era) (70.9%; 742/1047; p = 0.0365). There was no statistically significant difference between cases that had CHD codes at cardiac encounters (72.0%, 882/1225) and cases with codes at non-cardiac encounters (71.2%, 511/718) (p=0.7056). Cases with inpatient CHD codes had higher PPV (77.3%, 420/543) than cases with only outpatient CHD codes (62.5%, 581/930) (p<0.0001); however, PPV improved to 80.4% (333/414) when requiring at least 2 outpatient encounters with a CHD code separated by at least 30 days.

**Table 4.**
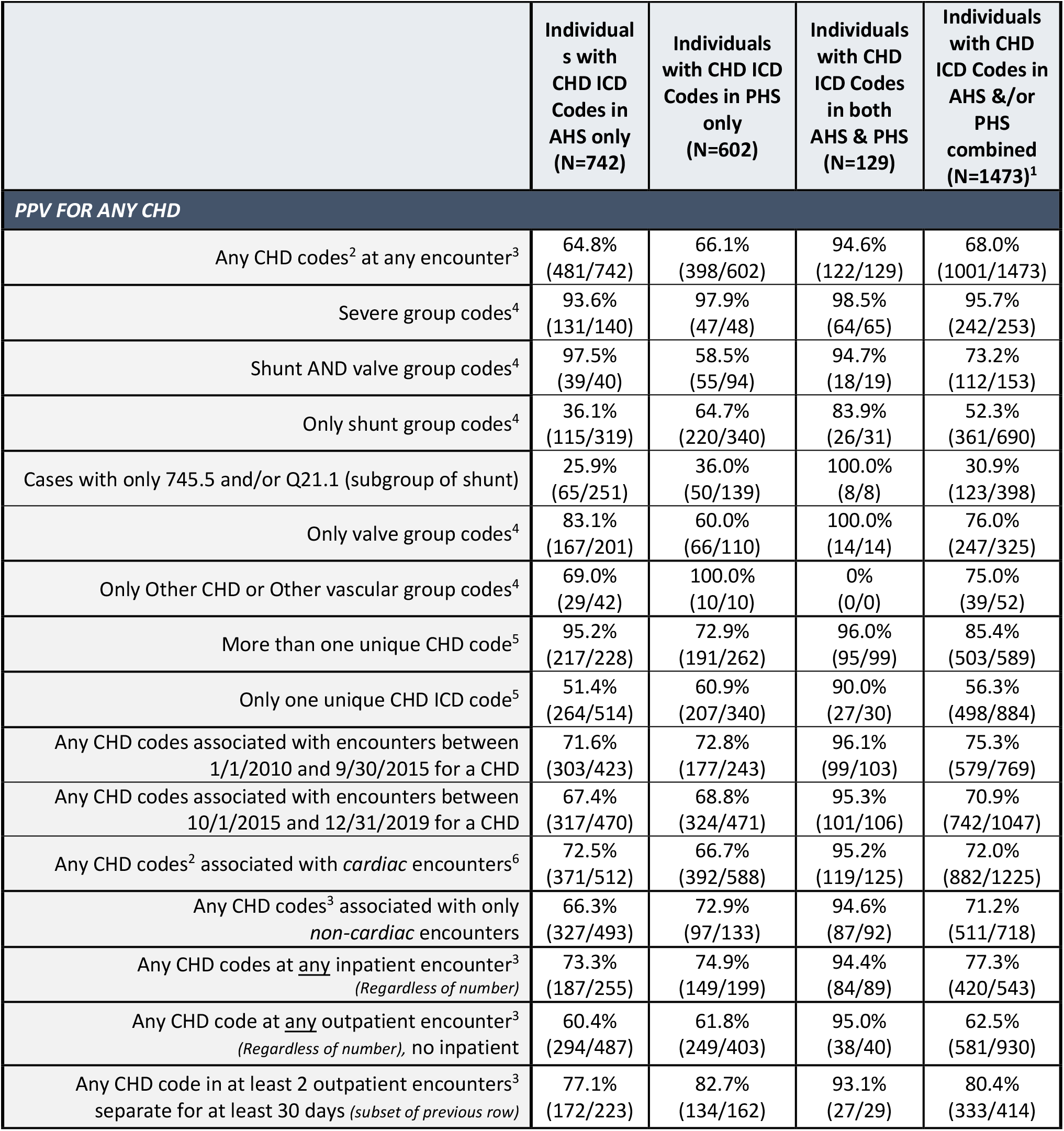
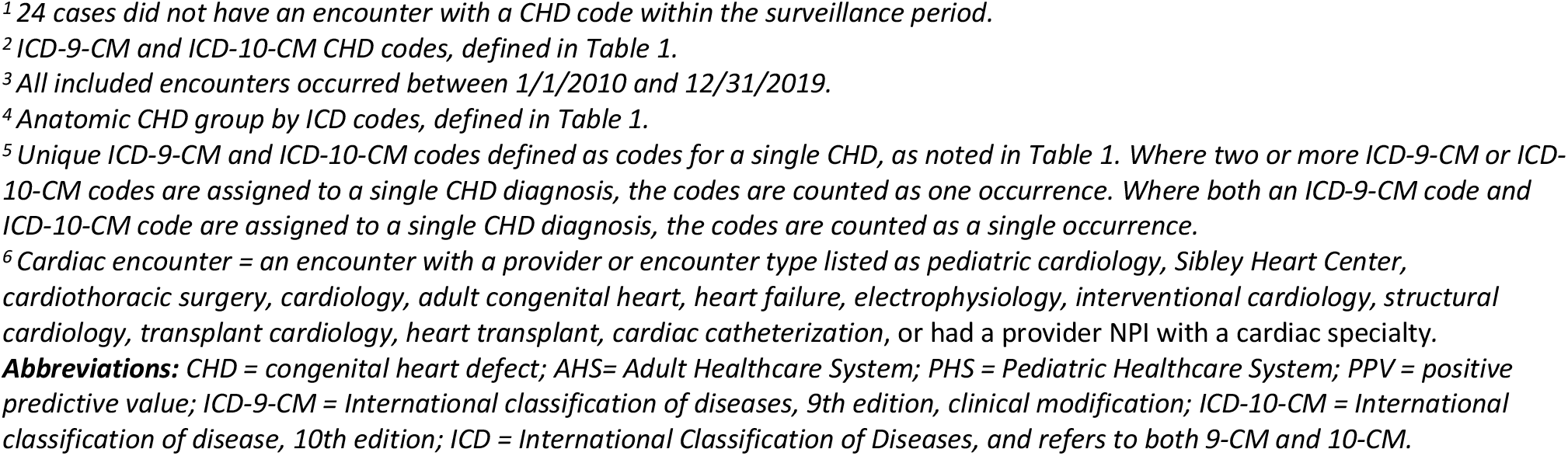
Positive Predictive Value (PPV) of CHD ICD-9-CM and ICD-10-CM Codes by Defect Characteristics for Identifying CHD Cases, Validated through Chart Abstraction in Electronic Health Records in Two Tertiary Healthcare Systems, and Overall.

|  | Individuals with CHD ICD Codes in AHS only (N=742) | Individuals with CHD ICD Codes in PHS only (N=602) | Individuals with CHD ICD Codes in both AHS & PHS (N=129) | Individuals with CHD ICD Codes in AHS &/or PHS combined (N=1473)<sup>1</sup> |
| --- | --- | --- | --- | --- |
| **PPV FOR ANY CHD** |  |  |  |  |
| Any CHD codes<sup>2</sup> at any encounter<sup>3</sup> | 64.8%<br>(481/742) | 66.1%<br>(398/602) | 94.6%<br>(122/129) | 68.0%<br>(1001/1473) |
| Severe group codes<sup>4</sup> | 93.6%<br>(131/140) | 97.9%<br>(47/48) | 98.5%<br>(64/65) | 95.7%<br>(242/253) |
| Shunt AND valve group codes<sup>4</sup> | 97.5%<br>(39/40) | 58.5%<br>(55/94) | 94.7%<br>(18/19) | 73.2%<br>(112/153) |
| Only shunt group codes<sup>4</sup> | 36.1%<br>(115/319) | 64.7%<br>(220/340) | 83.9%<br>(26/31) | 52.3%<br>(361/690) |
| Cases with only 745.5 and/or Q21.1 (subgroup of shunt) | 25.9%<br>(65/251) | 36.0%<br>(50/139) | 100.0%<br>(8/8) | 30.9%<br>(123/398) |
| Only valve group codes<sup>4</sup> | 83.1%<br>(167/201) | 60.0%<br>(66/110) | 100.0%<br>(14/14) | 76.0%<br>(247/325) |
| Only Other CHD or Other vascular group codes<sup>4</sup> | 69.0%<br>(29/42) | 100.0%<br>(10/10) | 0%<br>(0/0) | 75.0%<br>(39/52) |
| More than one unique CHD code<sup>5</sup> | 95.2%<br>(217/228) | 72.9%<br>(191/262) | 96.0%<br>(95/99) | 85.4%<br>(503/589) |
| Only one unique CHD ICD code<sup>5</sup> | 51.4%<br>(264/514) | 60.9%<br>(207/340) | 90.0%<br>(27/30) | 56.3%<br>(498/884) |
| Any CHD codes associated with encounters between 1/1/2010 and 9/30/2015 for a CHD | 71.6%<br>(303/423) | 72.8%<br>(177/243) | 96.1%<br>(99/103) | 75.3%<br>(579/769) |
| Any CHD codes associated with encounters between 10/1/2015 and 12/31/2019 for a CHD | 67.4%<br>(317/470) | 68.8%<br>(324/471) | 95.3%<br>(101/106) | 70.9%<br>(742/1047) |
| Any CHD codes<sup>2</sup> associated with *cardiac* encounters<sup>6</sup> | 72.5%<br>(371/512) | 66.7%<br>(392/588) | 95.2%<br>(119/125) | 72.0%<br>(882/1225) |
| Any CHD codes<sup>3</sup> associated with only *non-cardiac* encounters | 66.3%<br>(327/493) | 72.9%<br>(97/133) | 94.6%<br>(87/92) | 71.2%<br>(511/718) |
| Any CHD codes at <u>any</u> inpatient encounter<sup>3</sup><br>(Regardless of number) | 73.3%<br>(187/255) | 74.9%<br>(149/199) | 94.4%<br>(84/89) | 77.3%<br>(420/543) |
| Any CHD code at <u>any</u> outpatient encounter<sup>3</sup><br>(Regardless of number), no inpatient | 60.4%<br>(294/487) | 61.8%<br>(249/403) | 95.0%<br>(38/40) | 62.5%<br>(581/930) |
| Any CHD code in at least 2 outpatient encounters<sup>3</sup><br>separate for at least 30 days (*subset of previous row*) | 77.1%<br>(172/223) | 82.7%<br>(134/162) | 93.1%<br>(27/29) | 80.4%<br>(333/414) |

<sup>1</sup> 24 cases did not have an encounter with a CHD code within the surveillance period.
<sup>2</sup> ICD-9-CM and ICD-10-CM CHD codes, defined in Table 1.
<sup>3</sup> All included encounters occurred between 1/1/2010 and 12/31/2019.
<sup>4</sup> Anatomic CHD group by ICD codes, defined in Table 1.
<sup>5</sup> Unique ICD-9-CM and ICD-10-CM codes defined as codes for a single CHD, as noted in Table 1. Where two or more ICD-9-CM or ICD-10-CM codes are assigned to a single CHD diagnosis, the codes are counted as one occurrence. Where both an ICD-9-CM code and ICD-10-CM code are assigned to a single CHD diagnosis, the codes are counted as a single occurrence.
<sup>6</sup> Cardiac encounter = an encounter with a provider or encounter type listed as pediatric cardiology, Sibley Heart Center, cardiothoracic surgery, cardiology, adult congenital heart, heart failure, electrophysiology, interventional cardiology, structural cardiology, transplant cardiology, heart transplant, cardiac catheterization, or had a provider NPI with a cardiac specialty.
**Abbreviations:** CHD = congenital heart defect; AHS= Adult Healthcare System; PHS = Pediatric Healthcare System; PPV = positive predictive value; ICD-9-CM = International classification of diseases, 9th edition, clinical modification; ICD-10-CM = International classification of disease, 10th edition; ICD = International Classification of Diseases, and refers to both 9-CM and 10-CM.

Of the 1497 cases, 71.2% (185/260) of cases with severe codes had severe CHD, 26.5% (43/162) of cases with shunt and valve codes had shunt and valve CHD, 46.7% (328/703) of cases with shunt codes had shunt CHD, 71.2% (230/320) of cases with valve codes had valve CHD, and 71.2% (37/52) with “Other CHD and Other vascular” codes had other CHD/other vascular CHD. For those with shunt and valve codes, 32 of the 162 overall cases were confirmed to be shunt only, 40 were confirmed to be valve only, and 4 cases were neither shunt nor valve. Lastly, of the 46 women with documented pregnancies during 2010–2019, 33 had a confirmed CHD resulting in a PPV of 71.7% for CHD ICD codes documented during pregnancy.

## DISCUSSION

CHD represents a multitude of heart defects across a broad severity spectrum, so case ascertainment presents a challenge in efforts to assess prevalence and outcomes of those living with CHD. Administrative data can benefit CHD surveillance and research because it captures information on a large scale from many patients across multiple care settings; however, the data have variable and limited accuracy in detecting true CHD cases and may capture those with CHD-related diagnosis codes who do not actually have CHD.

Study results demonstrated overall modest, but variable, PPV of the group of ICD-9-CM and ICD-10-CM codes shown in Table 1 for detecting CHD cases after the exclusion of non-specific and non-cardiovascular ICD codes from the case definition. Features associated with higher PPV include identification of a case in more than one data source (PPV 94.6%), having CHD codes for severe CHD anatomy (PPV 95.8%), and having CHD codes associated with an inpatient encounter (77.3%) or 2 or more outpatient encounters (80.4%). However, some of these characteristics had relatively low prevalence in the data; fewer than 10% of cases were found in both data sources, and those with severe ICD codes made up only 17.4% of the dataset. Based on differences in distributions, TPs had more unique CHD ICD codes, unique CHD-related encounters, CHD coded cardiac encounters, and CHD coded non-cardiac encounters than FPs. A statistical difference in PPV was observed for cases coded using ICD-9-CM vs. ICD-10-CM, suggesting a small loss in accuracy with the transition to ICD-10-CM coding.

While PPV for CHD identified by ICD codes in administrative datasets could be increased by constraining the dataset to inpatient encounters, multiple unique CHD codes, multiple encounters with the CHD code, identification in more than one data source and/or to severe CHD codes, doing so could lead to the exclusion of many cases with true CHD and the resulting sample would not be representative of the larger CHD population. For example, requiring either an inpatient encounter with a CHD code or at least two outpatient encounters with a CHD code separated by 30 days for those without an inpatient encounter increased PPV from 68.0% to 80.4% in our sample, but resulted in a loss of 33.1% of TP cases from the dataset. Furthermore, such restrictions can bias the sample towards individuals with more severe CHD and/or more complicated clinical courses.

Similar to Rodriguez et al^11^, the current analysis confirmed a low PPV of 30.9% for ICD-9-CM and ICD-10-CM codes used for secundum ASD when occurring in the absence of other CHD codes, and 84.6% of FP were likely assigned this code because they had a PFO. Therefore, CHD studies may opt to exclude cases with isolated secundum ASD codes (745.5 or Q21.1) due to low PPV^24^, but they could be sacrificing more than 10% of true CHD cases from their sample. As of 10/01/2022, ASD and PFO have unique ICD-10-CM codes^25^, making future analyses of administrative data better able to identify true CHD cases and exclude those with PFO. However, as PFO did not have a unique ICD-10-CM code until recently, this issue cannot be resolved by code selection alone in studies including data prior to 10/01/2022, and thus will likely require algorithms or clinician review to differentiate cases with secundum ASD from those with PFO. Understanding features associated with TP cases captured with the secundum ASD codes and features associated with FP cases captured with secundum ASD codes would aid in improving data quality of CHD administrative datasets prior to 10/01/2022.

For the 33.8% of FP cases who did not have a PFO, we identified additional sources of miscoding, which included the use of CHD codes for other cardiac conditions that may be heritable, such Marfan or Brugada syndromes, heritable and metabolic cardiomyopathies, or non-congenital valvular heart disease, but that are not considered structural congenital heart defects. FP cases in women who are pregnant may result from CHD codes applied to the fetus rather than the pregnant women, creating another potential source of miscoding.

We further identified variable and limited accuracy in detecting correct anatomic severity group using ICD codes. Understanding features associated with TP versus FP cases and potential sources of miscoding is an initial step in developing algorithms to detect CHD more accurately in administrative records, prioritizing both PPV and sensitivity. Awareness of strengths and limitations of administrative data in characterizing the CHD population will help interpret study results. Several studies have found machine learning (ML) and natural language processing (NLP) to be promising for detecting heart failure cases, lupus cases, and symptoms in hemodialysis patients^26-29^. ML and NLP could potentially be applied to identify CHD cases more accurately in administrative datasets.^30^

## Limitations and Strengths

This ICD code validation study was strengthened by choosing specific ICD codes for the case definition of CHD rather than entire code groups, the use of ICD-9-CM and ICD-10-CM codes, high inter- and intra-observer reliability, and analysis of encounter-based features that are associated with TP and FP. Conversely, this study was limited by the fact that administrative data from tertiary referral center eMR are likely skewed towards more severe CHD given that individuals with severe CHD are most likely to seek care and, as such, any of our estimates not stratified by severity may be biased towards codes with a higher PPV. Tertiary referral centers may also code rare diseases with greater accuracy regardless of case severity. Therefore, the applicability of findings to other data sources, such as Medicaid data, may be limited. Additionally, ICD codes for CHD reflect native anatomy and thus we were unable to validate use of these codes for post-operative anatomy.

## Conclusion

Overall, ICD codes for CHD have modest PPV, which can be improved by requiring more than one unique CHD code, multiple encounters and/or data sources, or excluding codes with low PPV, but at the expense of lower sensitivity. Understanding the strengths and limitations of ICD CHD codes will help interpret research findings derived from administrative sources. Development of algorithms to detect CHD in administrative data may improve data quality.

## Data Availability

The data set contains PHI and cannot be shared publicly. The analysis has been replicated.

## AKNOWLEDGEMENTS

This study was supported in part by National Center for Advancing Translational Sciences of the National Institutes of Health under Award Number UL1TR002378 and the Medical Imaging, Informatics, and Artificial Intelligence Core (MIIAI), which is supported by the Department of Biomedical Informatics, Emory University School of Medicine. The content is solely the responsibility of the authors and does not necessarily represent the official views of the National Institutes of Health.

## eAppendix A

### Cardiac Imaging CPT Codes

#### Congenital Echocardiography codes

**93303** Transthoracic echocardiography for congenital cardiac anomalies; complete

**93304** Transthoracic echocardiography for congenital cardiac anomalies; follow-up or limited study

**93315** Transesophageal echocardiography for congenital cardiac anomalies; incl probe placement, image acquisition, interpretation, and report

**93316** Transesophageal echocardiography for congenital cardiac anomalies; placement of transesophageal probe only

**93317** Transesophageal echocardiography for congenital cardiac anomalies; image acquisition, interpretation & report only

#### Non-Congenital Echocardiography codes

**93306** Echocardiography, transthoracic, real-time with image documentation (2D), includes M-mode recording, when performed, complete, with spectral Doppler echocardiography, & with color flow Doppler echocardiography

**93307** Echocardiography, transthoracic, real-time with image documentation (2D), includes M-mode recording, when performed, complete, without spectral or color Doppler echocardiography

**93308** Echocardiography, transthoracic, real-time with image documentation (2D), includes M-mode recording, when performed, follow-up or limited study

**93312** Echocardiography, transesophageal, real-time with image documentation (2d) (with/without m-mode recording); incl probe placement, image acquisition, interpretation & report

**93313** Echocardiography, transesophageal, real-time with image documentation (2d) (with/without m-mode recording); placement of transesophageal probe only

**93314** Echocardiography, transesophageal, real-time with image documentation (2d) (with/without m-mode recording); image acquisition, interpretation & report only

**93318** Echocardiography, transesophageal (tee) for monitoring purposes, incl probe placement, real time 2-dimensional image acquisition & interpretation leading to ongoing (continuous) assessment of (dynamically changing) cardiac pumping function & to therapeutic measures on immediate time basis

**93320** Doppler echocardiography, pulsed wave and/or continuous wave with spectral display (list separately in addition to codes for echocardiographic imaging); complete

**93321** Doppler echocardiography, pulsed wave and/or continuous wave with spectral display (list separately in addition to codes for echocardiographic imaging); follow-up or limited study (list separately in addition to codes for echocardiographic imaging)

**93325** Doppler echocardiography color flow velocity mapping (list separately in addition to codes for echocardiography)

**C8925** Transesophageal echocardiography (tee) with/without contrast followed by with contrast, real time with image documentation (2d) (with/without m-mode recording); incl probe placement, image acquisition, interpretation & report

**C8926** Transesophageal echocardiography (tee) with/without contrast followed by with contrast, for congenital cardiac anomalies; incl probe placement, image acquisition, interpretation & report

**C8927** Transesophageal echocardiography (tee) with/without contrast followed by with contrast, for monitoring purposes, incl probe placement, real time 2-dimensional image acquisition & interpretation leading to ongoing (continuous) assessment of (dynamically changing) cardiac pumping function & to therapeutic measures on an immediate time basis

#### Cardiac Magnetic Resonance Imaging (MRI) codes

**71555** Magnetic Resonance Angiography (MRA) of chest

##### Cardiac MRI without gadolinium contrast

**75557** Cardiac MRI for morphology & function without contrast

**75558** Cardiac MRI for morphology & function without contrast; with flow/velocity quantification

**75559** Cardiac MRI for morphology & function without contrast; with stress imaging

**75560** Cardiac MRI for morphology & function without contrast; with flow/velocity quantification & stress

##### Cardiac MRI with gadolinium contrast

**75561** Cardiac MRI for morphology & function without contrast followed by contrast and further sequences

**75562** Cardiac MRI for morphology & function without contrast followed by contrast and further sequences; with flow/velocity quantification

**75563** Cardiac MRI for morphology & function without contrast followed by contrast and further sequences; with stress imaging

**75564** Cardiac MRI for morphology & function without contrast followed by contrast and further sequences; with flow/velocity quantification and stress

##### Cardiac MRI Add-on Flow Code

**75565** Cardiac MRI for velocity flow mapping

#### Cardiac Computed Tomography (CT) codes

**71725** CT angiography of the chest (used for looking for extracardiac anatomy such as pulmonary arteries, pulmonary veins, aortic arch, vascular rings)

**75571** Computed tomography, heart, without contrast, with quantitative evaluation of coronary calcium

**75572** Computed tomography, heart, with contrast, for evaluation of cardiac structure & morphology (incl 3D image postprocessing, assessment of cardiac function & evaluation of venous structures, if performed)

**75573** Computed tomography, heart, with contrast, for evaluation of cardiac structure & morphology in setting of congenital heart disease (incl 3D image postprocessing, assessment of LV cardiac function, RV structure & function & evaluation of venous structures, if performed)

**75574** Computed tomographic angiography, heart, coronary arteries & bypass grafts (when present), with contrast, incl 3D image postprocessing (incl evaluation of cardiac structure & morphology, assessment of cardiac function, & evaluation of venous structures, if performed)

